# Quantifying Static Postural Control Impairment after Stroke using Markerless Motion Capture

**DOI:** 10.64898/2026.10.03.26363941

**Authors:** Sachitha R.B.A. Wijekoon, Jereme B. Outerleys, Benjamin R. Ritsma, Kevin J. Deluzio, Vincent G. DePaul, Stephen H. Scott

## Abstract

Markerless motion capture offers a practical approach to biomechanical assessment in clinical populations outside of controlled laboratory environments. The purpose of this study was to evaluate the utility of markerless motion capture for quantifying static postural control impairment in individuals with stroke. Participants with subacute stroke (n=26) and healthy controls (n=171) performed a 30-second quiet standing task across three data collection sites in Kingston, Ontario, Canada. Video data were processed using Theia3D, and pelvis center of mass (COM) trajectories were extracted. Five discrete COM parameters were computed and transformed into Z-scores using normative models developed from the healthy population. An overall Task Score was calculated by aggregating parameter Z-scores. Forty-six percent of participants with stroke were identified as impaired on the task (Task Score > 95th percentile of the healthy distribution). At the parameter level, impairments were more prevalent in the mediolateral (ML) than anteroposterior (AP) direction (root-mean-square distance: 50% impaired ML vs. 12% impaired AP; mean velocity: 46% impaired ML vs. 35% impaired AP). Task scores and several task parameters demonstrated significant correlations with Berg Balance Scale scores (Spearman’s r = −0.64 to −0.81) and Functional Independence Measure Motor Sub-Scale scores (Spearman’s r = −0.62 to −0.77), with poorer performance associated with lower clinical scores. These findings demonstrate the utility of markerless motion capture for quantifying static postural control impairment following stroke. Future work should extend this framework to evaluate impairment in more dynamic postural control and gait tasks.

## 1. INTRODUCTION

Postural instability is a common impairment following stroke, contributing to increased fall risk, reduced independence, and diminished quality of life (Forster and Young, 1995; Garland et al., 2007; Schmid and Rittman, 2009). Given its strong predictive relationship with post-stroke mobility and independent living, postural control has become a major target of rehabilitation (Fukuda et al., 2025; van de Port et al., 2006).

Clinicians typically rely on ordinal rating scales to visually assess balance impairment and inform rehabilitation strategies (dos Santos et al., 2023; Sibley et al., 2013, 2011). For example, the Berg Balance Scale (BBS) is a common assessment that rates performance on 14 items using a scale of 0 (unable to perform) to 4 (able to perform), producing a total score reflecting overall balance ability and recommendations for functional mobility (e.g., gait aid use) (Berg et al., 1995). While these tools are simple to use and can have strong psychometric properties (Blum and Korner-Bitensky, 2008; Chinsongkram et al., 2016), ordinal rating scales often exhibit floor/ceiling effects and have limited sensitivity to subtle differences underlying impairment (Mancini and Horak, 2010). This is compounded by the inherent limitations of visual observation in clinical assessment (Williams et al., 2009). Addressing these limitations with more objective, sensitive assessment approaches may therefore better quantify post-stroke impairment and inform rehabilitation strategies.

Objective posturography based on kinematic and/or kinetic data is widely used in human movement analysis but has shown limited real-world clinical utility (Visser et al., 2008). This is largely explained by practical barriers facing existing technology. Optoelectronic marker-based motion capture systems require physical markers to be placed on the skin, a process which is both time-consuming and imposes strict clothing restrictions that may be impractical in clinical populations (Colyer et al., 2018; Wade et al., 2022). The need for specialized equipment and technical expertise further confine marker-based systems to laboratory environments. Force plates present fewer such barriers, although laboratory-grade systems still require specialized installation (Richmond et al., 2018). Portable alternatives have helped address this limitation (Lake et al., 2018; Richmond et al., 2018; Walsh et al., 2021); however, force plates are still limited in the breadth of functional tasks and movement information that can be captured.

Advances in computer vision have led to markerless motion capture solutions that address many of the practical barriers facing existing technology. These systems use deep-learning algorithms to estimate three-dimensional (3D) pose from single- or multi-camera setups (Wade et al., 2022). By eliminating the need for physical markers and strict clothing restrictions, these systems vastly reduce data collection times and participant burden, making them more feasible for integration outside of controlled laboratory environments (McGuirk et al., 2022; Riazati et al., 2022). Theia3D is a multi-camera markerless solution that has been extensively validated against marker-based systems primarily for gait analysis (Carvalho et al., 2025; Kanko et al., 2024, 2021; Seuthe et al., 2025; Wishaupt et al., 2024; Wren et al., 2023), but also for measuring postural control (Chaumeil et al., 2024; Federolf et al., 2025). Recently, (Majoni et al., 2026) reported strong correlations and low measurement error between Theia3D and marker-based estimates of the center of mass (COM) in individuals with stroke performing a series of balance tasks. This supports the validity of Theia3D for measuring postural control after stroke, although no work to date has evaluated its use for quantifying postural control impairment in this population.

A key unknown that remains unaddressed by prior validation work is the utility of Theia3D in realistic clinical environments. During real-world assessments, clinicians may need to position themselves near patients as a safety precaution, particularly for individuals with greater neurological or functional impairment. Their presence, however, creates a potential challenge to markerless pose estimation due to possible occlusions. To address this gap, we recently evaluated the ability of Theia3D to quantify static postural control in a cohort of healthy participants standing both alone and with a second person standing nearby to simulate clinical supervision (Wijekoon et al., 2026). We found strong agreement between Theia3D and force plate estimates of the COM in both conditions. Moderate to excellent test-retest reliability was also observed for several discrete COM parameters, most notably the 95% confidence ellipse area and mediolateral (ML) path length. These findings demonstrated that Theia3D could be used to assess postural control in clinical contexts, although the presence of supervision should be considered in study design, as it did have an impact on parameter reliability.

The objective of the present study was to extend this work to evaluate the utility of Theia3D for quantifying static postural control impairment in individuals with subacute stroke, an important period in stroke recovery marked by substantial change and improvement in functional ability (Kwakkel et al., 2006, 2004). We compared performance in individuals with stroke to normative models built from a large dataset of healthy controls, to better quantify impairment at the individual level (Scott et al., 2023). We also compared performance assessed with Theia3D to that assessed with traditional clinical assessment tools.

## 2. MATERIALS AND METHODS

### 2.1 Participants

Neurologically healthy control participants were recruited from Kingston, Ontario, through promotional flyers and word-of-mouth. Healthy control participants were included if they were ≥ 18 years of age and able to provide informed consent. They were excluded if they had any known neurological or musculoskeletal conditions impacting postural control. Participants who had experienced stroke were recruited through inpatient and outpatient rehabilitation programs at Providence Care Hospital, Kingston, Ontario. Participants with stroke were included if they were ≥ 18 years of age, in the subacute phase of stroke (7 days to 6 months), able to ambulate for short distances with or without an assistive device, and able to provide informed consent. They were excluded if they had any non-stroke neurological condition, neurodegenerative disease, or lower-limb injury impacting mobility or postural control. Although efforts were made to recruit participants with a first-ever stroke, those with a previous history of stroke were permitted to participate if they otherwise met all inclusion criteria. Height and weight information was obtained from clinical charts for participants with stroke and by self-report for healthy controls. All participants provided written informed consent, and ethics approval was given by the Queen’s University Health Sciences and Affiliated Teaching Hospital Research Ethics Board (DBMS-147-22).

### 2.2 Clinical Measures

Participants with stroke were assessed by a trained physiotherapist (PT) using several standardized clinical assessments. Postural control was assessed using the Berg Balance Scale (BBS), which evaluates 14 balance tasks on a scale of 0 (unable to perform) to 4 (able to perform), for a total score out of 56 (Berg et al., 1995). Stroke severity was assessed using the National Institutes of Health Stroke Scale (NIHSS), which assesses multiple neurological domains commonly affected by stroke for a total score ranging from 0 to 42, with higher scores indicating greater severity (Lyden, 2017). Disability in activities of daily living were characterized using both the Functional Independence Measure (FIM) and Modified Rankin Scale (mRS). We used the motor subscale of the FIM (FIM-M) which evaluates 13 items related to daily activity on a scale of 1 (total assistance) to 7 (complete independence), for a maximum score of 91 (Black et al., 1999). The mRS is a single-item scale that categorizes post-stroke disability from 0 (no symptoms) to 5 (severe disability) (van Swieten et al., 1988). The “affected” lower limb was further defined as the hemiparetic limb (commonly the contralateral side to the brain lesion). Motor impairment of the affected lower-limb was assessed using the lower extremity subscale of the Fugl-Meyer Assessment (FMA-LE), which evaluates 17 movements on a scale of 0 (cannot perform) to 2 (fully perform) for a maximum score of 34 (Fugl-Meyer et al., 1975). In cases where both sides were affected, FMA-LE scores were obtained for both lower limbs, and the lower score was recorded. Finally, both participants with stroke and healthy controls completed the EuroQol-5D-5L (EQ-5D-5L) to assess perceived quality of life (Golicki et al., 2015).

### 2.3 Motion Capture Setup

Data for this study were collected from three rooms across two clinical centers in Kingston, Ontario: Providence Care Hospital (PCH) and Hotel Dieu Hospital (HDH). Two rooms were used at PCH based on space availability, while the Human Mobility Research Laboratory was used at HDH. Each room varied slightly in size but were similarly configured using 8 Sony RX0 II cameras (Sony Corporation, Minato, Japan) arranged in an oval configuration surrounding the collection volume (see Supplementary Fig. S1 for diagrams of each setup). The 8 cameras were time-synchronized using camera control boxes (CCB-WD1, Sony Corporation, Minato, Japan) and recorded full HD video (1920 x 1080 pixels) at 60Hz and a shutter speed of 1/125. Calibration trials were recorded at the start of each data collection day using a chessboard of known dimensions.

### 2.4 Motion Capture Data Collection

Participants were recorded performing a battery of balance and gait tasks, with largely similar protocols across rooms (see Supplementary Table I for the full list of tasks). Participants with stroke wore their usual clothing, which generally consisted of comfortable, non-restrictive attire, and were not given specific clothing instructions. Healthy controls wore a range of everyday clothing but were given instructions to avoid clothing that was excessively baggy. A 30-second quiet standing task that was collected in all rooms was selected for analysis in the present study. During this task, participants were instructed to stand quietly with feet approximately hip-width apart (or in a self-selected comfortable stance for participants with stroke) (Fig. 1A-B). Only 30 seconds of quiet standing was recorded for kinematic analysis, however, participants with stroke continued to stand for a total of 2 minutes to allow concurrent clinical assessment of unsupported standing performance as part of the BBS. All stroke assessments were conducted under the supervision of a trained PT, who stood within one or two feet beside the participant as a safety precaution if deemed necessary. A chair was positioned behind participants with stroke as an additional safety precaution, which was replicated with the healthy controls to maintain consistency.

**Fig. 1.**
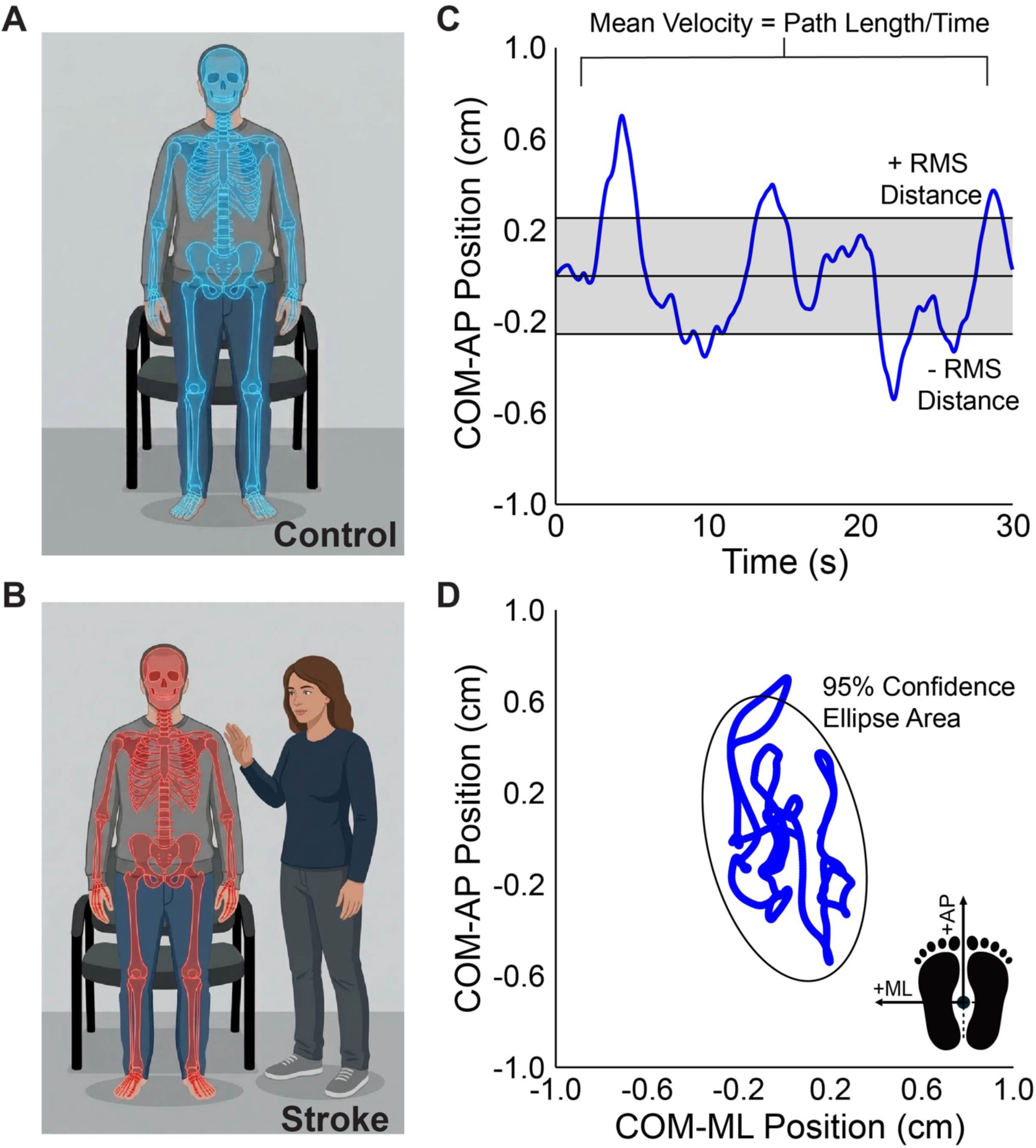
Experimental setup and discrete parameter visualization. A-B. Positioning of control participants (A) and participants with stroke (B) during the task. Participants stood with their hands by their sides and their feet roughly hip-width apart (or in the most comfortable position for participants with stroke). If deemed necessary, a PT stood one or two feet beside the participants with stroke. C. Parameter visualization: RMS distance and mean velocity. A representative healthy control participant’s COM-AP signal is shown. Mean velocity represents the path length (absolute sum of the point-to-point COM displacements) divided by trial time. RMS distance represents the RMS magnitude of the COM signal. As the COM signal was mean subtracted prior to calculation, this parameter is equivalent to the standard deviation of the signal. These parameters were also computed in the ML direction. D. Parameter visualization: 95% CE area. The same healthy control participant’s COM stabilogram is shown, representing sway along the AP-ML plane. The 95% CE area represents the area of the ellipse expected to enclose ∼95% of the points on the COM trajectory. The schematic on the bottom right visualizes the direction of positive AP and ML sway.

### 2.5 Data Processing

Video data were first processed in Theia3D (Theia Markerless Inc., Kingston, ON, v2026.1.4) to obtain 3D kinematics, using a 17-segment inverse kinematics model with three degrees-of-freedom at the hip, knee, and ankle. Kinematic data were exported in c3d file format for further processing in Visual3D (HAS-Motion Inc., Kingston ON, v2026.01.1). The COM of the pelvis segment was filtered in Visual3D using a fourth-order, zero phase-lag low-pass Butterworth filter with a 1Hz cut-off frequency. The COM signal was then exported to ASCII text files for analysis in R (R Core Team, 2023, v4.3.1).

### 2.6 Postural Control Parameters

Each participant’s anteroposterior (AP) and ML COM signals were mean subtracted prior to analysis.

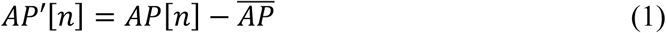

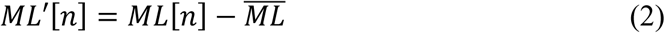

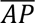 and 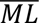 represent the mean of the AP and ML COM, respectively, and *AP*′[*n*] and *ML*′[*n*] represent the mean-subtracted signals at each time point n. From these signals, three postural sway parameters – root-mean-square (RMS) distance, mean velocity, and 95% confidence ellipse (CE) area – were calculated to characterize static postural control, derived from (Prieto et al., 1996). RMS distance and mean velocity were computed separately for the AP and ML directions, yielding five discrete parameters in total. In the following equations, N represents the total number of data points in the trial.

1. **RMS Distance:** The RMS magnitude of the COM signal. This is equivalent to the standard deviation of the signal, as both the AP and ML COM were mean subtracted prior to calculation.

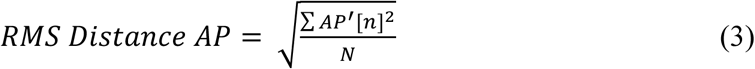

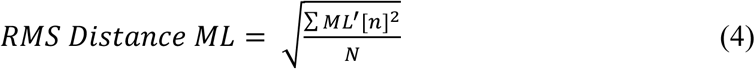
2. **Mean Velocity:** The sum of the absolute point-to-point COM displacements divided by the total trial time, t.

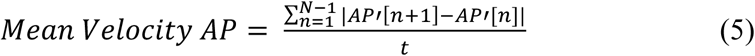

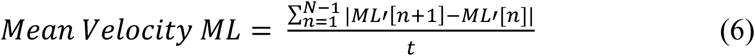
3. **95% CE Area:** The area of the ellipse enclosing approximately 95% of the points on the COM trajectory in the AP-ML plane.

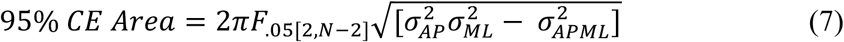 *F*_.05[2,*N*–2]_ is the critical value at the 95% confidence level with 2 and N-2 degrees of freedom. *σ_AP_* and *σ_ML_* represent the standard deviation of the AP and ML COM, respectively, while *σ_APML_* represents the covariance between the AP and ML time series.

These parameters were selected based on our previous findings of moderate to strong test-retest reliability and/or robustness to the presence of a second person (Wijekoon et al., 2026). Mean velocity was used instead of path length, as it is more commonly reported in the postural control literature (Lafond et al., 2004; Lin et al., 2008; Moghadam et al., 2011). These parameters are visualized in Fig. 1C-D. None of the parameters showed strong associations with anthropometric variables (height, weight, or base of support) (Supplementary Fig. S2) and therefore no normalization was applied.

### 2.7 Normative Modelling

Normative models of healthy performance were developed for each parameter, following the protocol used for assessments of the upper limb with the Kinarm Robotic Platform (Scott et al., 2023). For the detailed process, see the Kinarm Standard Tasks Summary (https://kinarm.com/download/kst-summary-analysis-version-3-11/). Briefly, raw parameter distributions were transformed into normal distributions using Box-Cox transformations, with transformed values then fit using weighted linear regression to account for age, sex, and the presence of a second person. The presence of a second person was incorporated as a covariate using the “Accompanied” data from our previous study, where healthy controls performed quiet standing with a second person standing nearby (Wijekoon et al., 2026). For these participants, first visit data from both the “Accompanied” and “Unaccompanied” (regular quiet standing) task were used for normative modeling. This process resulted in parameters expressed as standardized Z-scores (units of standard deviation). Z-scores beyond ±3.29 (1 in 1000) were removed as outliers, and the normalization step was repeated. Data renormalization and outlier removal were repeated a total of three times.

In addition to Z-scores for each parameter, overall Task Scores were generated. Specifically, parameter Z-scores were first transformed such that best performance = 0 and higher values indicated poorer performance. Transformed Z-scores were then aggregated using root-sum-squares, and the result was normalized using Box-Cox transformation following the same procedure as the parameter Z-scores. Participants with Z-Tasks Scores greater than 3.29 (1 in 1000) were removed as outliers, with data renormalization and outlier removal repeated up to three times. Task Scores were finally generated by transforming the result such that best performance = 0 and higher values indicated poorer performance.

Importantly, raw parameter values for participants with stroke were converted into Z-scores using the coefficients derived from the healthy models. This allowed impairment in stroke to be quantified relative to the expected range of the healthy population, accounting for age, sex, and the presence of a second person during assessment. Specifically, impairment in participants with stroke was defined as Z-scores > 1.65, or the 95^th^ percentile of the healthy control distribution. Task Scores for participants with stroke were similarly computed using the healthy models, with impairment defined as Task Scores > 1.96 (>95^th^ percentile of healthy controls).

### 2.8 Statistical Analysis

Parameter Z-scores and Task Scores were further compared to clinical outcome measures using Spearman rank correlations (r). Statistical significance was set at p < 0.05, with Bonferroni correction applied to adjust for multiple comparisons. Correlation strength was interpreted as negligible (r ≤ 0.30), low (0.31 ≤ r ≤ 0.50), moderate (0.51 ≤ r ≤ 0.70), high (0.71 ≤ r ≤ 0.90), and very high (r ≥ 0.91) (Mukaka, 2012).

## 3. RESULTS

### 3.1 Participant Demographics

A total of 177 healthy control participants and 28 participants with stroke participated in this study. Of these, 5 healthy controls and 2 participants with stroke were excluded due to improper task execution (e.g., voluntary movement during quiet standing). One additional healthy control was removed as an outlier during normative model development. The final analysis therefore included 171 healthy controls and 26 participants with stroke. Participant demographic and clinical characteristics are summarized in Table I (see Supplementary Table II for demographics split by collection room).

**Table 1.** Participant Demographic.

| Characteristic | Control (n=171) | Stroke (n=26) |
| --- | --- | --- |
| Demographic characteristics |  |  |
| Age [Mean (range)] | 42 (18-92) | 67.3 (41-90) |
| Male sex [n] | 53/118 | 19/7 |
| Gender identity [M/W/GD] | 53/116/2 | 19/7 |
| Stroke characteristics |  |  |
| Stroke Type [I/H] | - | 21/5 |
| Mean days post-stroke [SD] | - | 36 (28) |
| Affected Side [R/L/B] | - | 9/12/5 |
| Stroke Location [C/Sc/Br/Cb/<br>Br+Cb/C+Cb] | - | 11/10/5 |
| Clinical characteristics |  |  |
| BBS [Mean (SD)] | - | 17/5/1/1/1/1 |
| NIHSS [Mean (SD)] | - | 2.1* (1.8) |
| FIM-M [Mean (SD)] | - | 78 (14) |
| mRS [Mean (SD)] | - | 2.9 (0.56) |
| FMA-LE Affected Side***<br>[Mean (SD)] | - | 28** (2.6) |
| EQ-5D-5L Index [Mean (SD)] | - | 41 (11) |
| EQ-5D-5L VAS [Mean (SD)] | 0.92 (0.046) | 0.79 (0.13) |

Abbreviations: M/W/GD, Man/Woman/Gender Diverse; I/H, Ischemic/Hemorrhagic; SD, Standard Deviation; R/L/B, Right Side/Left Side/Both Sides; C/Sc/Br/Cb/Br+Cb/C+Cb, Cortex/Sub-Cortex/Brainstem/Cerebellum/ Brainstem + Cerebellum/Cortex + Cerebellum; BBS, Berg Balance Scale; NIHSS, National Institute of Health Stroke Scale; FIM-M, Functional Independence Measure – Motor; mRS, Modified Rankin Scale; FMA-LE, Fugl-Meyer Assessment – Lower Extremity; VAS, Visual Analogue Scale. \*n=23, \*\*n=25 (due to time constraints with some patients, certain clinical scales were not collected). \*\*\*For participants affected on both sides, the worse FMA-LE score was taken.

All participants with stroke were in the subacute phase (7 days to 6 months post-stroke), with most participants assessed within 1-month post-stroke (n = 17). Groups differed significantly in age (Wilcoxon Rank-Sum Test, p < 0.0001) and sex (Fisher’s Exact Test, p < 0.001), however, these variables were accounted for in normative modeling.

### 3.2 Representative Participant COM Signals

Representative COM stabilograms for a healthy control participant and a participant with stroke are shown in Fig. 2. Panel A shows a healthy male participant in his 80s, whose stabilogram reveals a greater range of AP relative to ML COM sway. Panel B shows a male participant with stroke also in his 80s (16 days post-stroke, BBS = 41, FIM-M = 83, FMA (affected) = 30, mRS = 3, NIHSS = 1) who was classified as impaired (Task Score = 2.56). For this participant, the range of AP sway was comparable to the healthy control, but the range of ML sway was much greater.

**Figure 2.**
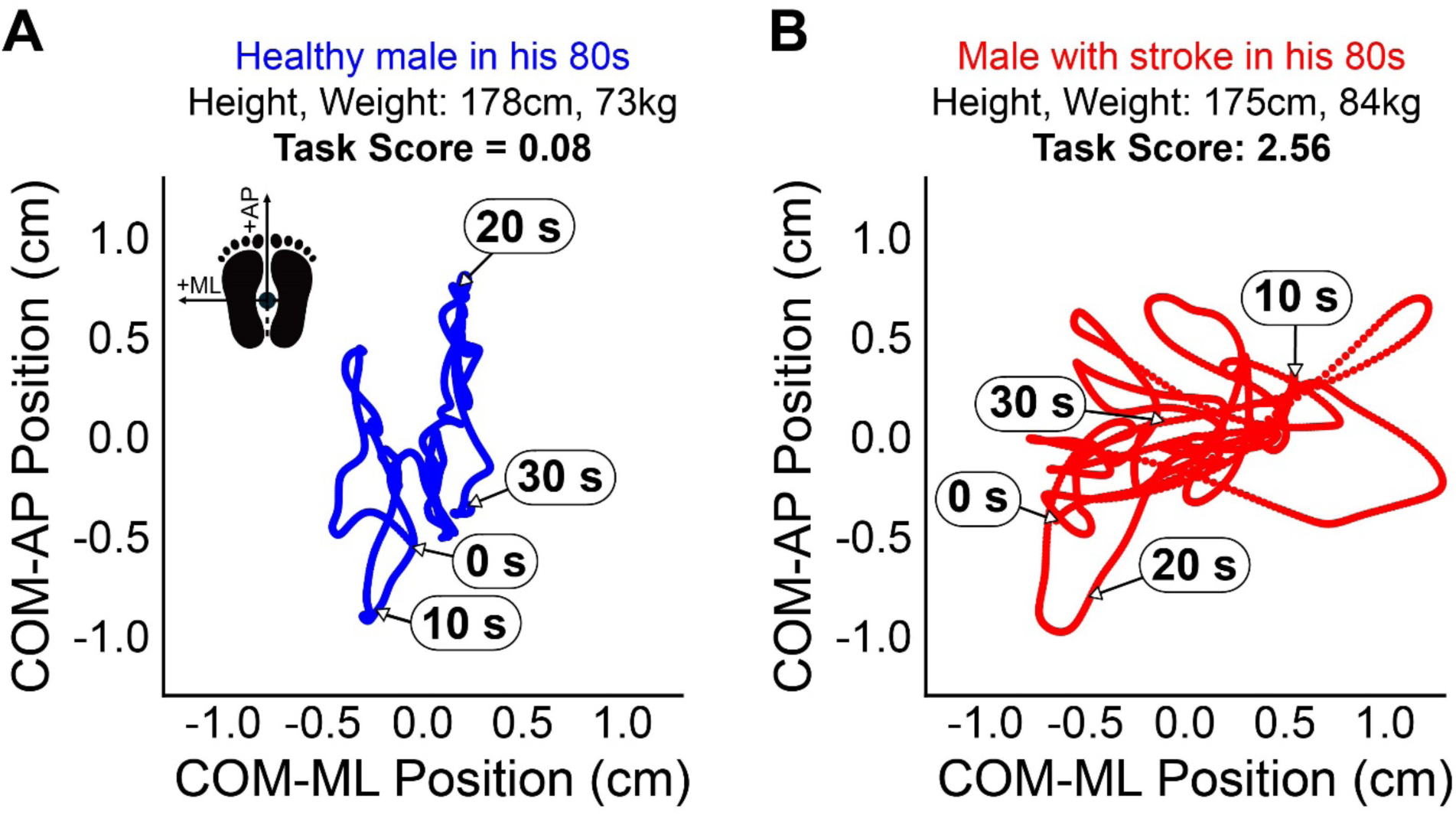
Representative healthy control and stroke participant COM stabilograms. Example COM stabilograms for a healthy control (A) and a participant with stroke classified as impaired (Task Score > 1.96). The y-axis represents the mean subtracted COM-AP signal, and the x-axis represents the mean subtracted COM-ML signal. The schematic on the healthy control stabilogram visualizes the directions of positive sway in the AP and ML directions. Timestamps are included on each stabilogram.

### 3.3 Parameter- and Task-Level Impairments

Fig. 3 illustrates cumulative sums of parameter and task performance for healthy controls and participants with stroke. Just under half (46%) of the participants with stroke were identified as impaired at the task level. At the parameter level, ML parameters identified more participants as impaired than AP parameters, most notably for RMS distance (50% impaired ML vs. 12% impaired AP) but also mean velocity (46% impaired ML vs. 35% impaired AP). The 95% CE area, which encompasses both AP and ML sway, classified 35% of participants with stroke as impaired.

**Figure 3.**
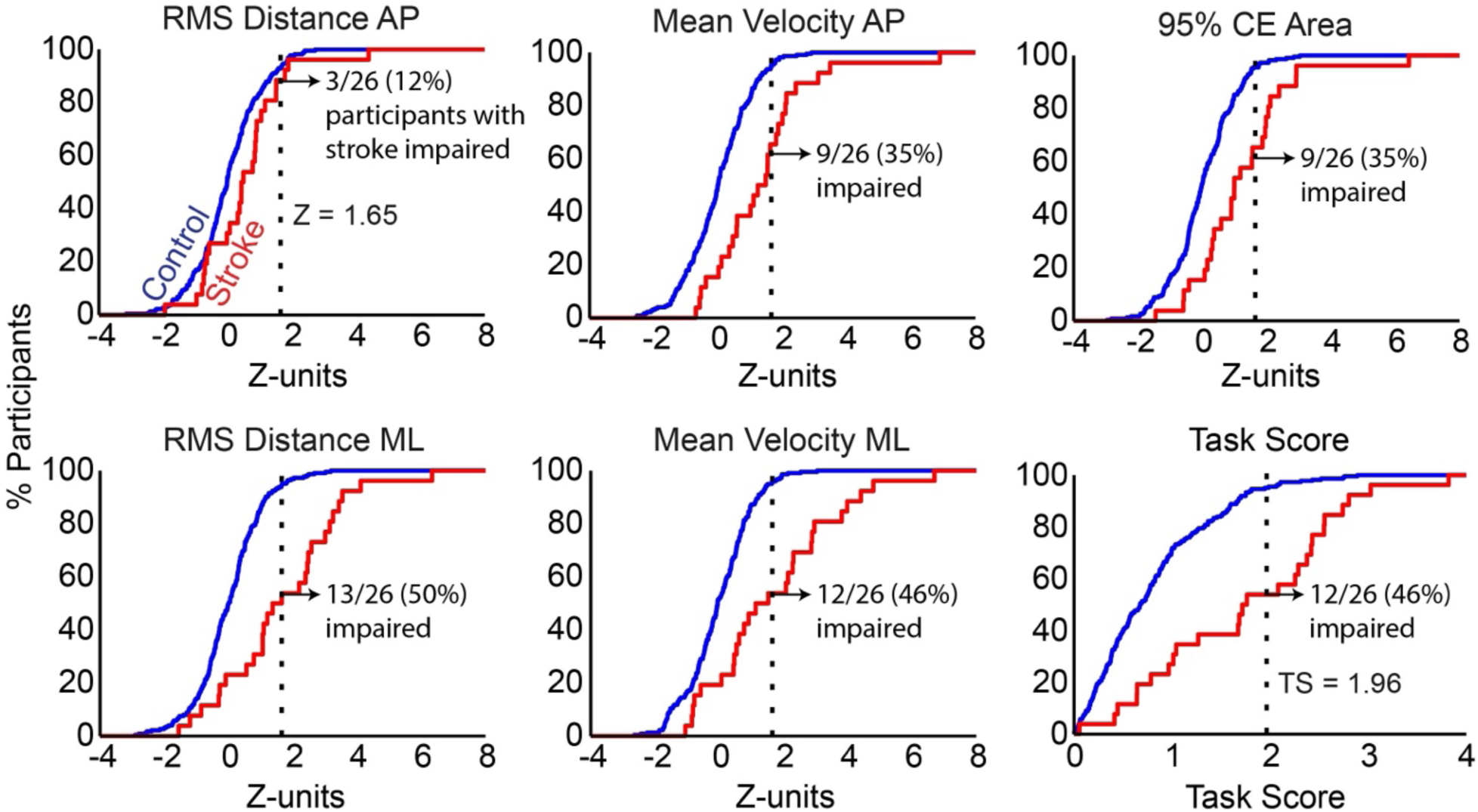
Cumulative sums of performance at parameter and task level. Cumulative sums for each task parameter and Task Score (TS) were compared between control participants (blue lines) and participants with stroke (red lines). The dotted vertical line at Z = 1.65 for each discrete parameter represents the 95^th^ percentile of control performance and was used as the threshold for impairment. A similar vertical dotted line at TS = 1.96 represents the same threshold for Task Score. The count and percentage of participants with stroke with performance beyond these thresholds, and therefore considered impaired, is indicated on each plot.

The directionality of impairments is further illustrated in Fig. 4. Panel A compares patterns of impairment direction separately across RMS distance and mean velocity parameters. A similar number of participants were identified as impaired by each parameter (13 for RMS distance, 14 for mean velocity), but the directional pattern of these impairments differed. For RMS distance, impairments were largely isolated to the ML direction, with 10 participants identified with isolated ML impairments and 3 with impairments in both directions. Impairments were more distributed for mean velocity, with 7 participants identified with impairments in both directions, 5 with isolated ML impairments, and 2 with isolated AP impairments.

**Figure 4.**
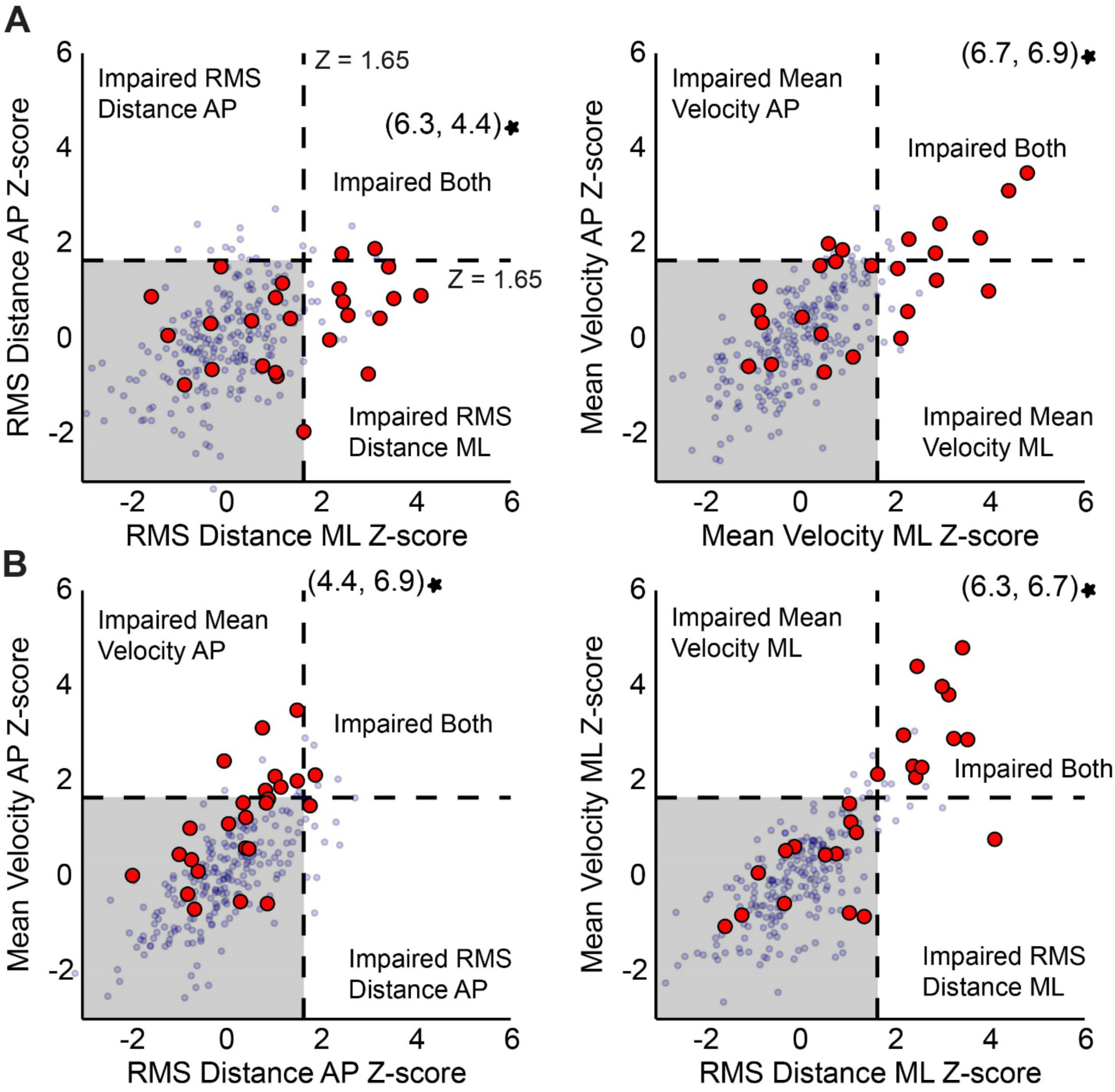
Comparison of impairments across directions and parameters. Impairments are compared across directions for each parameter (A) and across parameters for each direction (B). Impairments are compared across directions for each parameter (A) and across parameters for each direction (B). Red dots represent participants with stroke, whereas small blue dots represent healthy controls. The dotted horizontal and vertical lines at Z = 1.65 mark the impairment thresholds in both the AP and ML direction, enclosing the shaded gray area (bottom left quadrant). One participant had Z-scores beyond the plotted range for RMS distance ML, mean velocity AP, and mean velocity ML, and is indicated by a star on the relevant plots.

Panel B compares patterns of impairment separately between AP parameters and between ML parameters. AP impairments showed little coupling between RMS distance and mean velocity, with 7 participants identified with impairments only in AP mean velocity, 2 with impairments in both AP parameters, and 1 with impairment only in RMS distance AP. In contrast, ML impairments were mostly coupled between parameters, with 12 participants identified as impaired in both RMS distance ML and mean velocity ML, and 1 with impairment only in RMS distance ML.

### 3.4 Clinical Correlations

Of the five clinical measures assessed, only the BBS and FIM-M scores showed significant correlations with any task parameter or Task Score (Fig. 5). High negative correlations were observed between both clinical measures and RMS distance ML (BBS: r = − 0.81, p < 0.0001; FIM-M: r = −0.77, p < 0.001) and 95% CE area (BBS: r = −0.74, p < 0.001; FIM-M: r = −0.72, p < 0.001). Task Score and mean velocity ML were highly correlated with BBS Score (Task Score: r = −0.78, p < 0.0001; mean velocity ML: r = −0.71, p < 0.01) and moderately correlated with FIM-M score (Task Score: r = −0.70, p < 0.01; mean velocity ML: r = −0.62, p < 0.05). Mean velocity AP was also moderately correlated with BBS score (r = −0.64, p < 0.05). Scatter plots depicting clinical correlations are provided in Supplementary Fig. S5 – S9.

**Figure 5.**
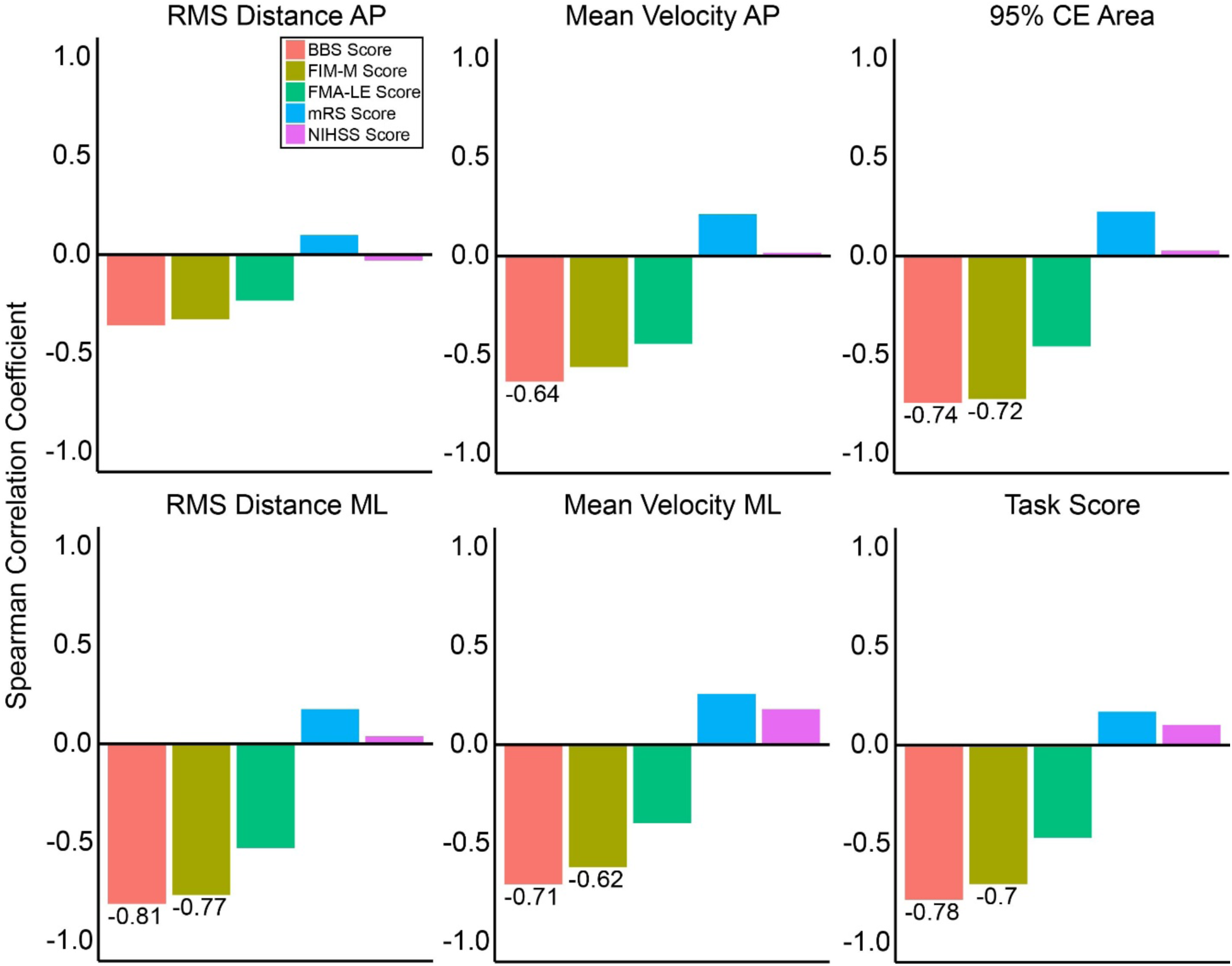
Spearman’s correlations between clinical measures and task parameters/Task Score. Each bar shows the strength of correlation, with the correlation coefficient indicated for significant correlations (p < 0.05).

## 4. DISCUSSION

We investigated the utility of Theia3D markerless motion capture for quantifying static postural control impairment in individuals with stroke during a 30-second quiet standing task. Using our normative modeling framework, the Task Score identified 46% of participants with stroke as impaired relative to the healthy population. Parameter-level analyses further revealed a greater prevalence of ML compared to AP impairments. Finally, several significant negative correlations were observed between clinical measures and both parameter Z-scores and Task Scores.

Quantifying performance in individuals with stroke relative to the healthy distribution represents a step toward identifying patient-specific impairments. Many studies continue to draw clinical insights by comparing means and/or standard deviations between age- and sex-matched patient and healthy cohorts (Halmi et al., 2020; Wang et al., 2019; Yu et al., 2008). While these comparisons can provide valuable group-level insights, individual-level inferences cannot be made by comparing performance to a healthy mean. Normative modeling addresses this limitation by estimating expected performance based on relevant characteristics of the healthy population, allowing individuals with stroke to be assessed relative to what is expected from healthy individuals with similar characteristics (Scott et al., 2023). For example, the performance of a male with stroke in his 60s can be evaluated relative to what is expected from a typical healthy male in his 60s. An additional benefit of our normative modeling approach is that parameters measured on different scales are standardized into Z-scores, permitting direct comparisons across parameters. Further aggregation of parameter Z-scores into a composite Task Score provides an overall representation of task performance, which may offer a clinically useful summary of the extent of impairment across multiple aspects of postural control.

The observed pattern of AP vs. ML sway impairment can be interpreted through the dynamics of postural control in quiet standing. During typical bipedal stance, the inverted pendulum model describes AP balance as being predominantly controlled through ankle plantar/dorsiflexor activity, whereas ML balance is controlled primarily through a hip load/unload mechanism involving the hip ab/adductors (Winter, 1995; Winter et al., 1996). AP control is dominant during typical stance, characterized by relatively synchronous activity between the limbs. Conversely, ML control relies more on coordinated shifts in weight distribution between the limbs. This may be particularly relevant following stroke, as hemiparesis often results in asymmetric weight distribution between the paretic and non-paretic limb, which has been associated with increased ML sway (Marigold and Eng, 2006). More broadly, previous studies comparing healthy and stroke populations have reported greater ML sway following stroke, while differences in AP sway may be more closely associated with age-related changes (Dault et al., 2003; Yu et al., 2008). Therefore, the greater prevalence of ML impairment observed in this study may reflect the particular vulnerability of ML postural control to stroke-related motor deficits.

Quiet standing performance as measured by markerless motion capture demonstrated several significant correlations with clinical measures. Worse performance, particularly in the ML direction, was correlated with worse performance on the BBS and FIM-M. The relationship with BBS performance was expected, given that the quiet standing task was taken directly from the BBS and performed concurrently with the clinical assessment. Nevertheless, the strength of these correlations across task parameters is notable, as it suggests similar performance information could be obtained from the single 30-second quiet standing task as could be from the entire 14-item balance scale. The strong correlations with FIM-M are less direct, as the FIM-M assesses functional independence across a broader range of motor activities. This relationship may reflect the importance of postural stability to mobility and activities of daily living more broadly following stroke (Geurts et al., 2005). Of the clinical measures that were not significantly correlated with quiet standing performance, the lack of association with the FMA-LE was particularly notable given its direct assessment of lower-limb motor impairment. However, lower-limb impairment may not necessarily translate to greater standing instability as quantified by the COM, as individuals could potentially compensate through their non-paretic limb or other postural strategies.

Markerless motion capture is a rapidly advancing technology that is making objective biomechanical data collection more practical and accessible. The present study provides an example of this, with data from individuals with stroke collected directly in a hospital setting, with no specific clothing restrictions imposed and PT supervision provided when needed, leading to partial occlusions in some camera views. Still, increased accessibility should not be conflated with the absence of practical considerations. Factors such as external occlusions from clinicians or self-occlusion caused by clothing can affect pose estimation and increase the potential for tracking errors. Markerless systems such as Theia3D do benefit from ongoing improvements to pose-estimation models and algorithms, which can allow previously collected data to be reprocessed with potentially improved tracking. Nevertheless, improved algorithms cannot fully compensate for poor-quality input data. This consideration becomes increasingly important as Theia3D is used beyond controlled laboratory settings to address novel questions in clinical populations and challenging environments. The data collection principles recommended by the manufacturer should continue to be carefully considered and adapted as necessary (Theia, n.d.). For example, in this study, clothing restrictions for participants with stroke were deemed impractical, but the camera configuration was optimized and PTs positioned themselves to limit occlusion across multiple cameras when possible. Ultimately, researchers should determine the most appropriate setup based on the specific population, environment, and intended application of their study.

This study has limitations. Constraints in patient recruitment resulted in a relatively small sample of individuals with stroke, precluding meaningful subgroup analyses of different clinical characteristics. Considering the substantial heterogeneity in motor impairment following stroke, which can be influenced by factors such as stroke type, lesion location, and pre-existing comorbidities, these characteristics may have contributed to the variability in impairment observed (Alawieh et al., 2018; Geurts et al., 2005). Data were also collected across multiple sites, and site was not included as a covariate in the normative models. However, supplementary analyses did not identify significant differences in COM parameters between sites (see Supplementary Tables III and IV). This study also only evaluated static postural control through a simple 30-second quiet standing task. Postural control involves the integration of sensory information from somatosensory, visual, and vestibular systems, which can be differentially challenged depending on the nature of the task (Horak, 2006). Future work should evaluate a broader range of postural control tasks to capture impairments that may not be apparent during quiet standing alone. Additionally, four participants with stroke only had 28-29 seconds of usable data, although this is not expected to systematically bias parameter estimates. Finally, we used the COM of the pelvis as a surrogate measure for the total-body COM as it was more robust to occlusions. This choice was supported by our previous study, which demonstrated strong correlation and agreement between pelvis and total-body COM during quiet standing (Wijekoon et al., 2026).

In summary, this study demonstrated that Theia3D is able to quantify static postural control impairment in individuals with stroke. Impairments were more prevalent in the ML compared to the AP direction. Task performance was strongly correlated with relevant clinical measures, particularly for ML parameters. Future work should extend this framework to more complex tasks and longitudinal assessments to provide a more comprehensive characterization of postural control impairment and recovery following stroke.

## Data Availability

The data used in the present study are available upon reasonable request to the corresponding author. Clinical data are not available to the public as the information could compromise the privacy of the participants with stroke.

## Acknowledgements

We would like to thank K. Moore, M.J. Demers, and L. Samis for their help with participant recruitment and data collection. We would also like to thank the members of the Human Mobility Research Laboratory, for allowing us to use their research space. This research was supported by the Ontario Research Fund (ORF-RE-09-112), the Natural Sciences and Engineering Research Council of Canada (RGPIN-2024-04486), and The Hilary and Galen Weston Foundation. The funding sources were not involved in the completion of this study. The authors would also like to thank the participants in this study for giving their time.

