## Supplementary Material for "Quantifying Static Postural Control Impairment after Stroke using Markerless Motion Capture"

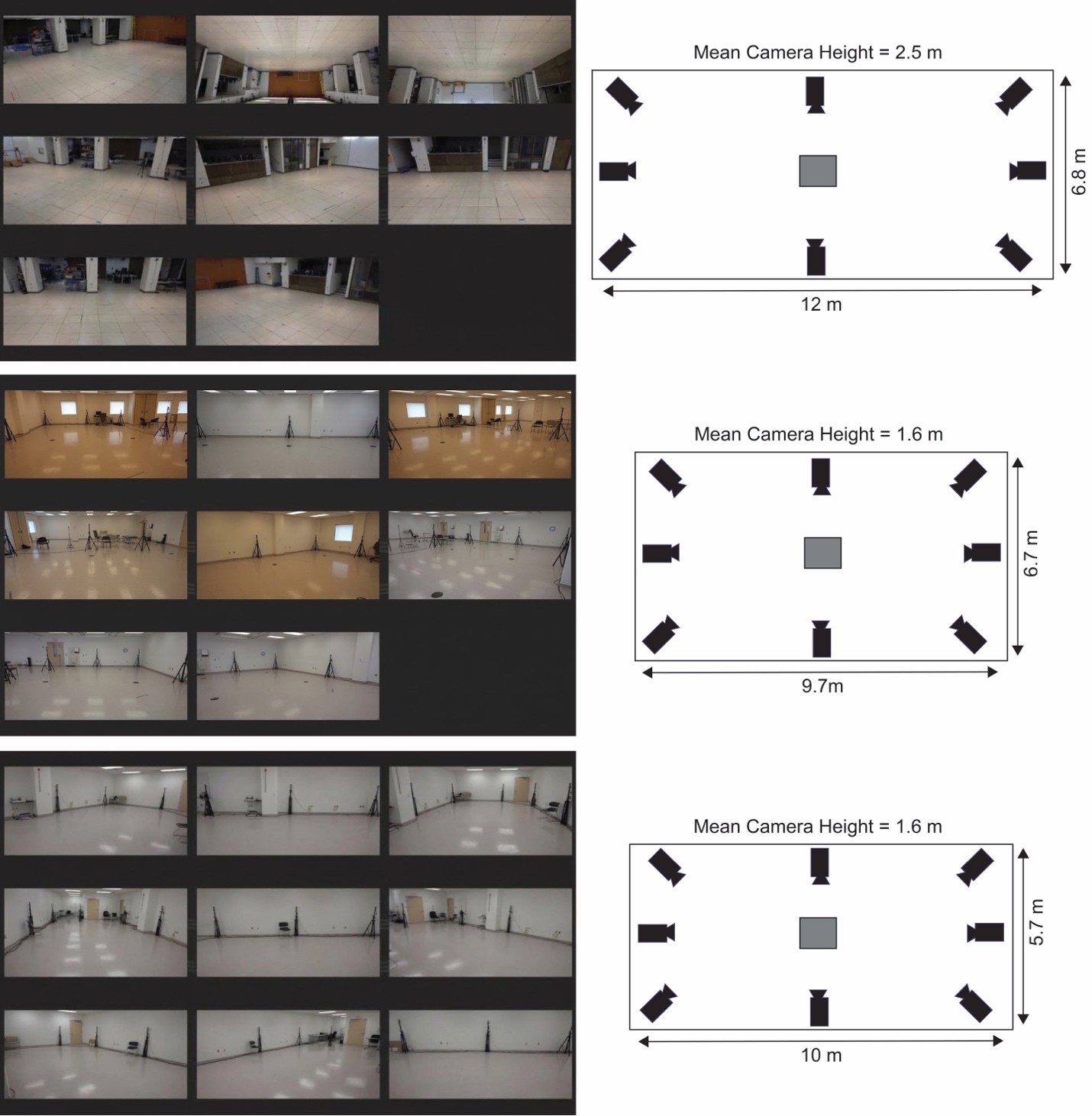
**Supplementary Figure S1. Experimental setup across collection rooms (top = HDH, middle = PCH_1, bottom = PCH_2).** Panels in the left column show camera views in each room. Panels in the right column show room dimensions with approximate camera locations and mean camera heights.

**Supplementary Table I. List of recorded tasks**

| **HDH*** | **PCH_1** | **PCH_2** |
| --- | --- | --- |
| - **1-min walk:** participants walked at a comfortable pace back and forth between two pieces of tape for 1 minute. - **Timed Up and Go:** participants stood up from a chair, walked forward 3 metres, turned around, walked back to the chair, and sat back down. This task was also performed at a comfortable walking pace. - **Quiet Stand:** participants stood with their feet roughly hip-width apart and arms by their side for 30 seconds. - **Quiet Stand Eyes Closed:** participants stood with their feet roughly hip-width apart, arms by their side, and eyes closed for 10 seconds. - **Quiet Stand Feet Together:** participants stood with feet together and arms by their side for 60 seconds.   *Note: participants completed tasks both alone, and with a second person walking or standing beside them, simulating clinical supervision. For more details, see Wijekoon et al., 2026. | - **1-min walk:** same as HDH. - **5-metre walk test:** participants walked at a comfortable pace between two pieces of tape spaced 5 metres apart. - **Timed Up and Go:** same as HDH. - **Berg Balance Scale:** participants completed a series of 14 static and dynamic balance tasks. Tasks were simultaneously recorded and scored by a PT for individuals with stroke. - **Fugl-Meyer Upper/Lower Extremity Coordination:** participants were recorded completing part D and part F of the Fugl-Meyer Upper and Lower Extremity assessments, respectively. The tasks were performed seated as opposed to supine to allow for better markerless tracking. | - **1-min walk:** same as HDH. - **5-metre walk test:** same as PCH_1. - **Timed Up and Go:** same as HDH. - **Berg Balance Scale:** same as PCH_1. - **Fugl-Meyer Upper/Lower Extremity Coordination:** same as PCH_1. |

**Normalization Test**

Spearman’s correlation between each discrete parameter and the following anthropometric variables were computed: height, weight, and base-of-support area (Supplementary Fig. S2). Base-of-support area was computed using six foot landmarks defined in Visual3D: left and right heel, left and right fifth metatarsal, and left and right big toe. For each frame, the convex hull of these six points was calculated using the convhulln() function from the geometry package in R, which fits the smallest convex polygon enclosing all landmark coordinates. The enclosed area of this polygon was averaged across all frames to produce a single average base-of-support area.

Outliers ±3 SD of the mean for each parameter x anthropometric variable combination (42/3765 or ~0.01% of total observations) were removed to avoid skewing correlations and improve visualization. Bonferroni correction was applied across all correlation tests to correct for multiple comparisons.

Several significant correlations were observed, most notably with all parameters having significant positive correlations with weight. However, correlation strength ranged from negligible to low for all significant correlations. As such, parameters were not normalized by anthropometric variables.

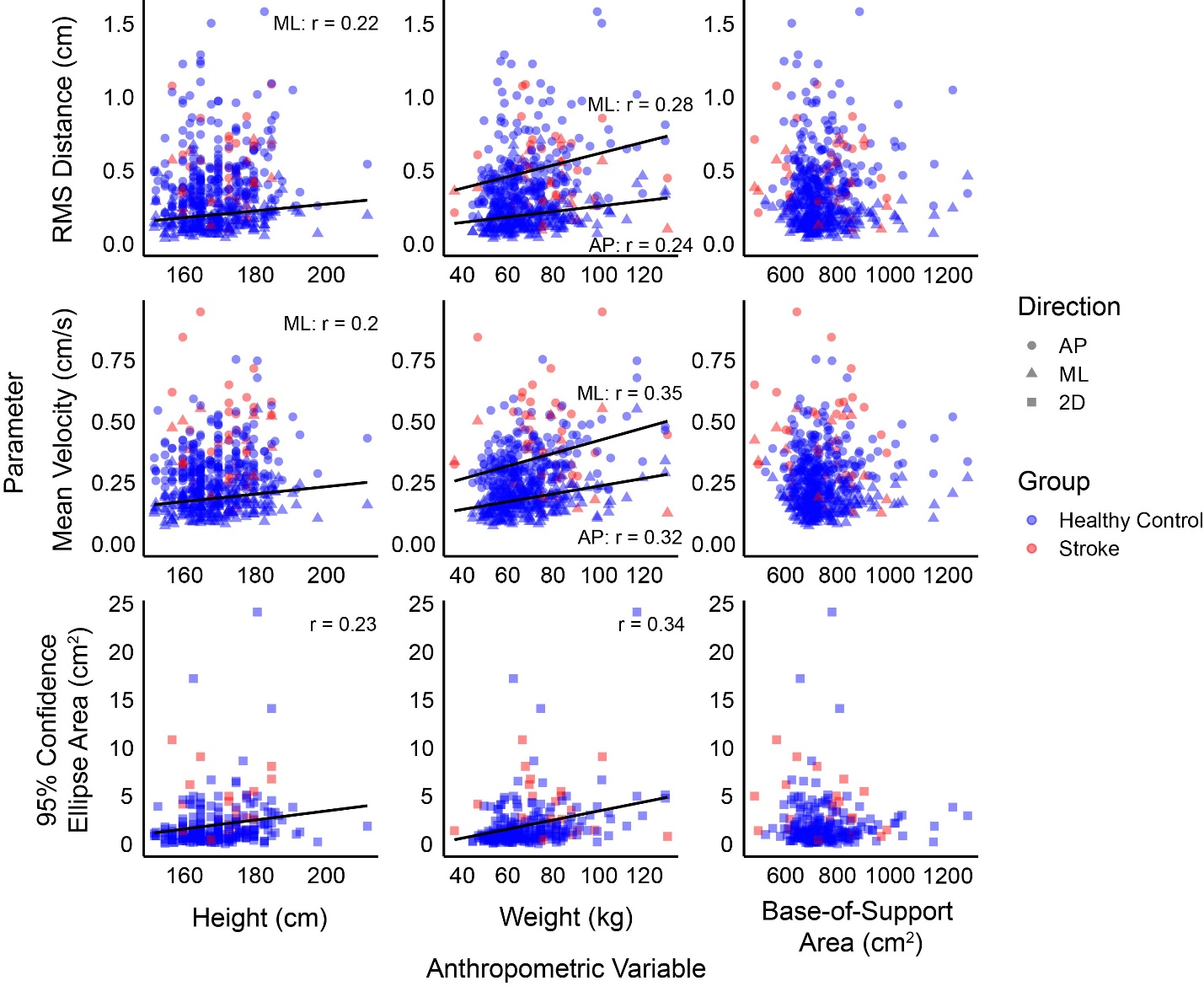
**Figure S2. Correlation between COM parameters and anthropometric variables.** RMS distance (top row), mean velocity (middle row), and 95% confidence ellipse area (bottom row) values are plotted on the y-axis, against values for height (left column), weight (middle column), and base-of-support (right column) on the x-axis. Points on each plot are separated by signal direction (AP = circles, ML = triangles, 2D AP-ML plane = squares). Blue shapes represent healthy participants while red shapes represent participants with stroke. Outliers ±3 SD of the mean for each parameter x anthropometric variable combination (42/3765 or ~0.01% of total observations) were removed to avoid skewing correlations and improve visualization. The Spearman’s correlation coefficient is indicated for each relevant signal direction, if significant (p < 0.05).

**Supplementary Table II. Demographics information split by collection site**

| **Characteristic** | **Control HDH**  **(n=79)** | **Control PCH_1 (n=68)** | **Control PCH_2 (n=24)** | **Stroke PCH_1 (n=20)** | **Stroke PCH_2 (n=6)** |
| --- | --- | --- | --- | --- | --- |
| Demographic characteristics | | | | | |
| Age [Mean (range)] | 25 (18-68) | 57 (22-92) | 57 (20-89) | 67 (41-90) | 69 (43-87) |
| Male sex [n] | 21 | 22 | 10 | 15 | 4 |
| Gender identity [M/W/GD] | 21/56/1 | 22/46 | 10/14 | 15/5 | 4/2 |
| Stroke characteristics | | | | | |
| Stroke Type [I/H] | - | - | - | 18/2 | 3/3 |
| Mean days post-stroke [SD] | - | - | - | 36 (28) | 35 (12) |
| Affected Side [R/L/B] | - | - | - | 8/8/4 | 4/1/1 |
| Stroke Location [C/Sc/Br/Cb/  Br+Cb/C+Cb] | - | - | - | 13/4/0/1/1/1 | 4/1/1/0/0/0 |
| Clinical characteristics | | | | | |
| BBS [Mean (SD)] | - | - | - | 40 (12) | 45 (11) |
| NIHSS [Mean (SD)] | - | - | - | 2.2* (1.9) | 1.5** (1.3) |
| FIM-M [Mean (SD)] | - | - | - | 78 (12) | 78 (20) |
| mRS [Mean (SD)] | - | - | - | 3 (0.51) | 2.8 (0.75) |
| FMA-LE Affected Side****  [Mean (SD)] | - | - | - | 28*** (2.3) | 29 (3.8) |
| EQ-5D-5L Index [Mean (SD)] | 0.93 (0.041) | 0.93 (0.037) | 0.9 (0.075) | 0.79 (0.13) | 0.79 (0.16) |
| EQ-5D-5L VAS [Mean (SD)] | 83 (10) | 88 (7.6) | 85 (8.9) | 68 (19) | 69 (12) |

Abbreviations: M/W/GD, Man/Woman/Gender Diverse; I/H, Ischemic/Hemorrhagic; SD, Standard Deviation; R/L/B, Right Side/Left Side/Both Sides; C/Sc/Br/Cb/Br+Cb/C+Cb, Cortex/Sub-Cortex/Brainstem/Cerebellum/ Brainstem + Cerebellum/Cortex + Cerebellum; BBS, Berg Balance Scale; NIHSS, National Institute of Health Stroke Scale; FIM-M, Functional Independence Measure – Motor; mRS, Modified Rankin Scale; FMA-LE, Fugl-Meyer Assessment – Lower Extremity; VAS, Visual Analogue Scale. *n=19, **n=4, ***n=19 (due to time constraints with some patients, certain clinical scales were not collected). ****For participants affected on both sides, the worse FMA-LE score was taken.

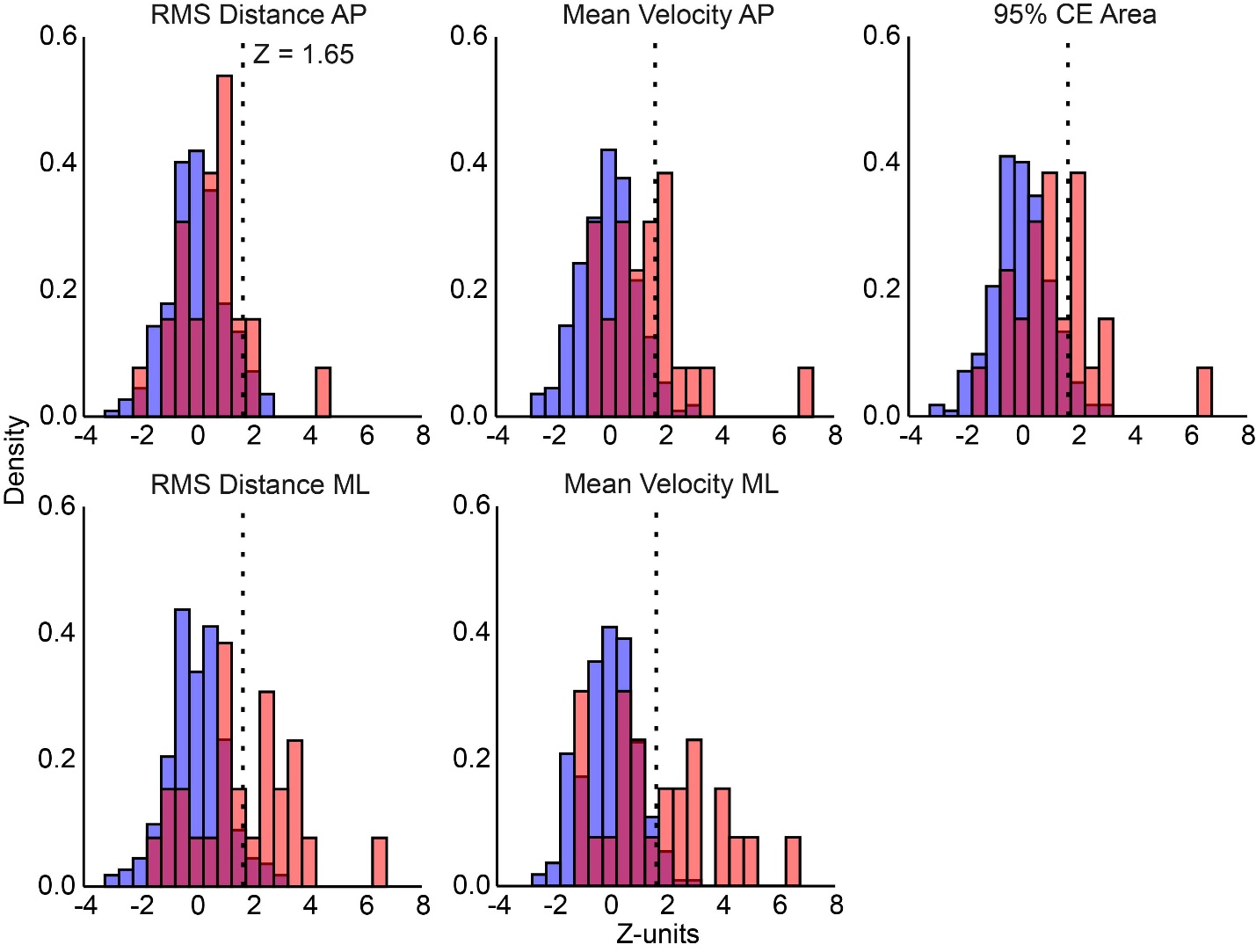
**Figure S3.** Histograms of Z-scores for each parameter, scaled to probability density. Blue and red bars represent healthy control and stroke distributions, respectively. The dotted vertical line represents Z = 1.65, or the 95th percentile of healthy performance. Histogram bin width was set to 0.5.

**Testing for site effects**

As our normative model did not account for site effects, we assessed for any significant differences across sites in both our healthy control participants and participants with stroke (Supplementary Tables III and IV). Healthy control data were collected across three sites, while stroke data were collected across two sites. A one-way ANOVA was therefore used to assess site effects in the healthy control group, with data collected at HDH with a second person nearby excluded to satisfy the independence of observations assumption. A Wilcoxon rank-sum test was used for the stroke group. P-values were adjusted using Bonferroni correction across all five tests per group. Following correction, no significant site effects were observed for any parameter in either group, suggesting that Z-scores derived from the normative model are robust to site-related variability. It should be noted however that the small number of stroke patients at PCH_2 (n = 6) limits the statistical power of this comparison, and the absence of a significant site effect in the stroke group should be interpreted with caution.

**Supplementary Table III. Site effects in healthy controls (one-way ANOVA)**

| Parameter | F | df | p | p (adjusted) |
| --- | --- | --- | --- | --- |
| RMS Distance AP | 2.89 | 2, 168 | 0.058 | 0.292 |
| RMS Distance ML | 0.14 | 2, 168 | 0.871 | 1.000 |
| Mean Velocity AP | 0.43 | 2, 167 | 0.653 | 1.000 |
| Mean Velocity ML | 3.73 | 2, 165 | 0.026 | 0.131 |
| 95% CE Area | 1.17 | 2, 168 | 0.314 | 1.000 |

Abbreviations: F, F statistics; df, degrees of freedom. Note: p-values adjusted using Bonferroni correction.

**Supplementary Table IV. Site effects in participants with stroke (Wilcoxon rank-sum)**

| Parameter | W | p | p (adjusted) |
| --- | --- | --- | --- |
| RMS Distance AP | 75.0 | 0.387 | 1.000 |
| RMS Distance ML | 84.0 | 0.157 | 0.783 |
| Mean Velocity AP | 82.0 | 0.196 | 0.981 |
| Mean Velocity ML | 80.0 | 0.242 | 1.000 |
| 95% CE Area | 86.0 | 0.123 | 0.615 |

Abbreviations: W, W statistic. Note: p-values adjusted using Bonferroni correction.

**Receiver Operator Characteristic (ROC) Analysis**

To further evaluate the discriminative ability of each parameter and the composite Task Score, ROC analysis was performed (Supplementary Fig. S4). ROC curves describe the trade-off between sensitivity (true positive rate) and 1 - specificity (false positive rate) across all possible Z-score thresholds, reflecting each parameter's ability to discriminate between healthy control participants and participants with stroke. Discriminative performance was quantified using the area under the curve (AUC), with values closer to 1 indicating better discrimination.

ROC analyses were conducted for each parameter using the roc() function from the pROC package in R. Ninety-five percent confidence intervals for AUC were estimated from 1000 bootstrap resamples.

Overall, AUC values were relatively close (between 0.76 and 0.79) across all parameters except for RMS Distance AP.

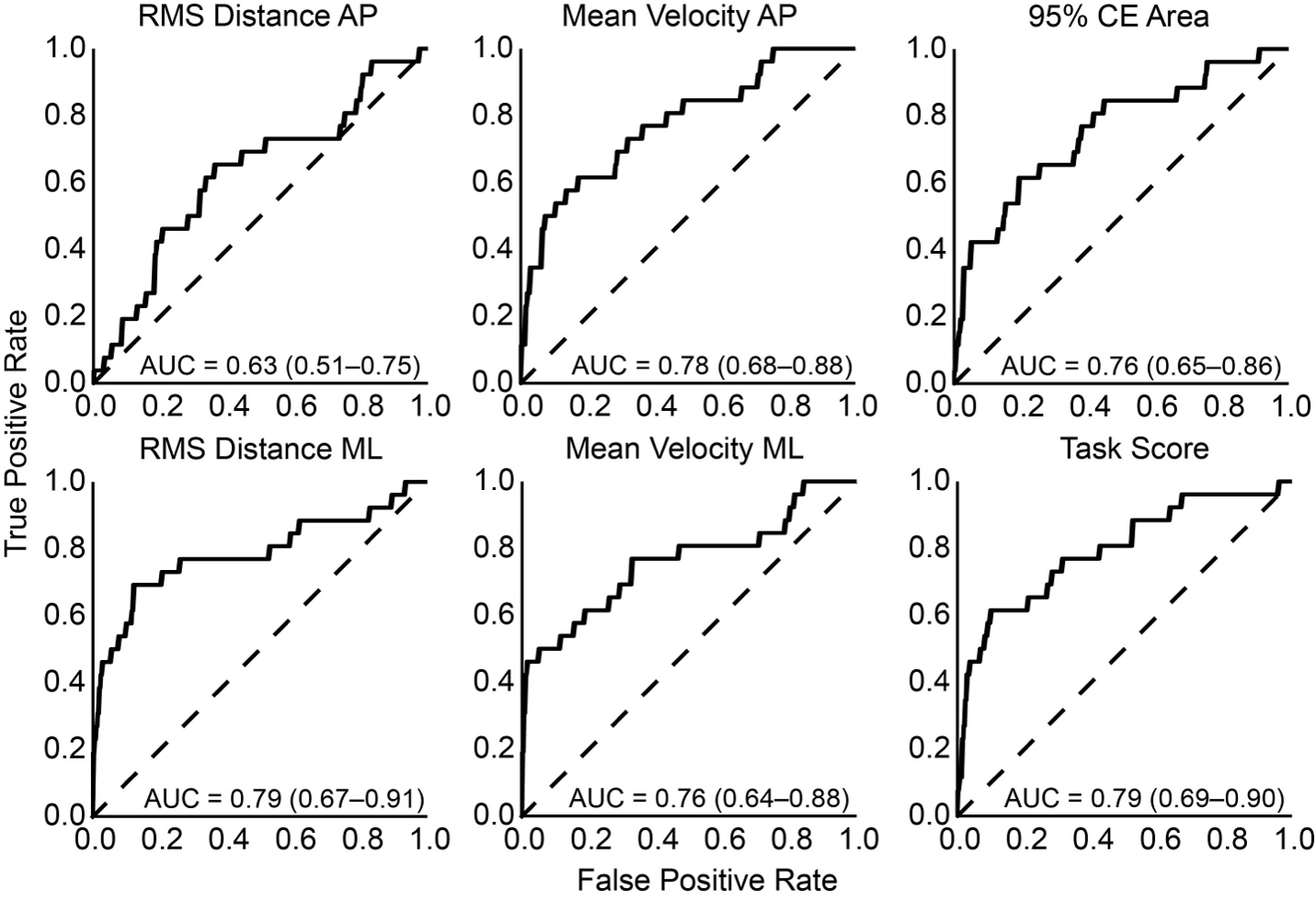
**Figure S4. ROC curves for each COM parameter and Task Score.** AUC values and corresponding 95% confidence intervals (1000 bootstrap resamples) are reported for each parameter. The dashed identity line represents chance-level discrimination (AUC = 0.50).

**Comparison of Quiet Standing Performance and Additional Clinical Measures**

Spearman rank correlations were computed between each task parameter and the following clinical measures: Berg Balance Scale (BBS) score, Functional Independence Measure Motor (FIM-M) score, the Fugl-Meyer Lower Extremity score, Modified Rankin Scale (mRS) score, and the National Institutes of Health Stroke Scale (NIHSS) score. These correlations were summarized in Figure 5 of the main manuscript. Individual scatter plots depicting each correlation in greater detail can be found in Supplementary Fig. S5 – S9.

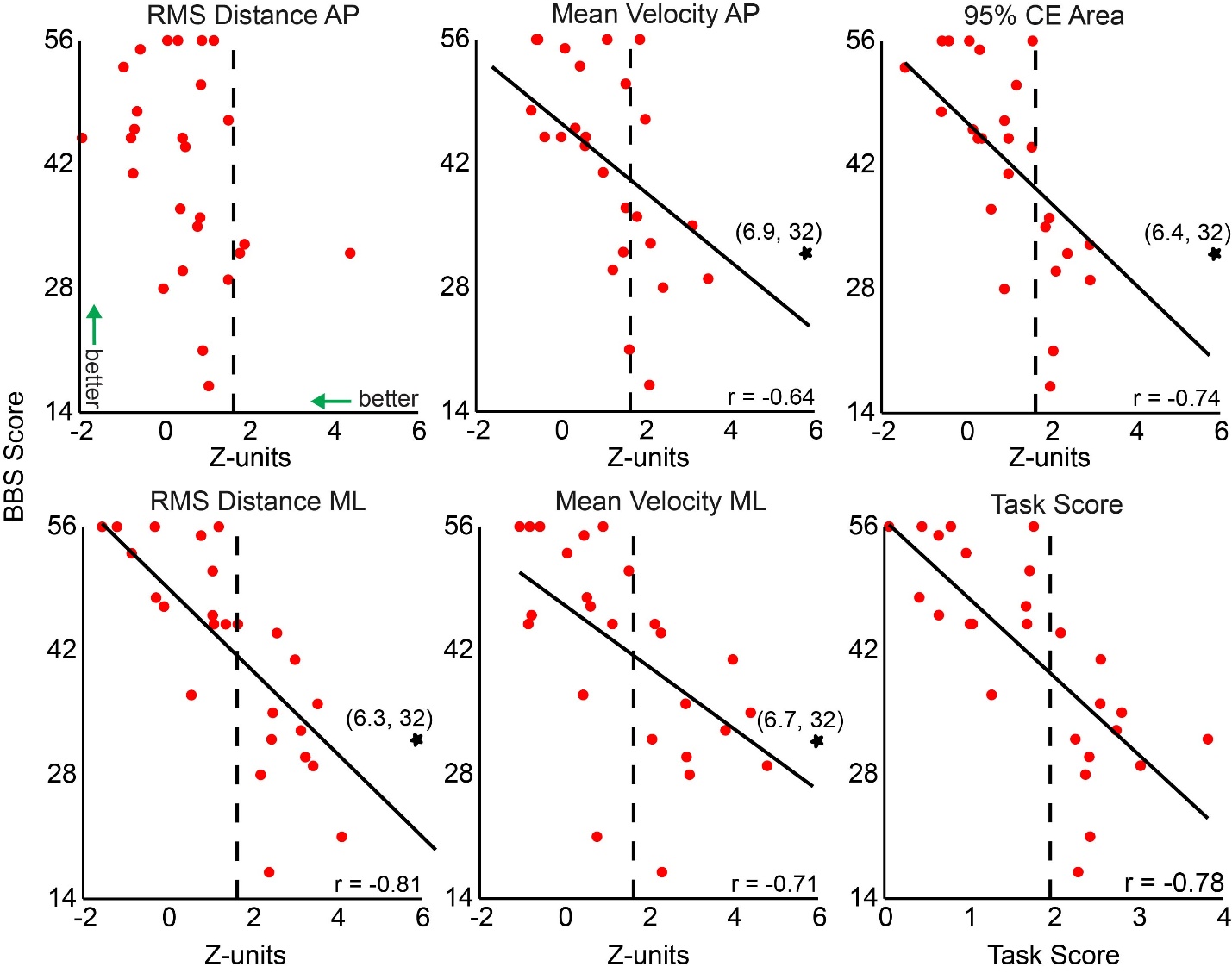

**Figure S5. Correlations between BBS scores and task parameters/Task Score.** Each red dot represents a participant with stroke. The dotted vertical line at Z = 1.64 for each discrete parameter represents the 95^th^ percentile of control performance, which was the threshold of impairment. A similar vertical dotted line at Task Score = 1.96 represents the same threshold for Task Score. One subject had Z-scores beyond the plotted range for several parameters and is indicated by a star on the relevant plots.

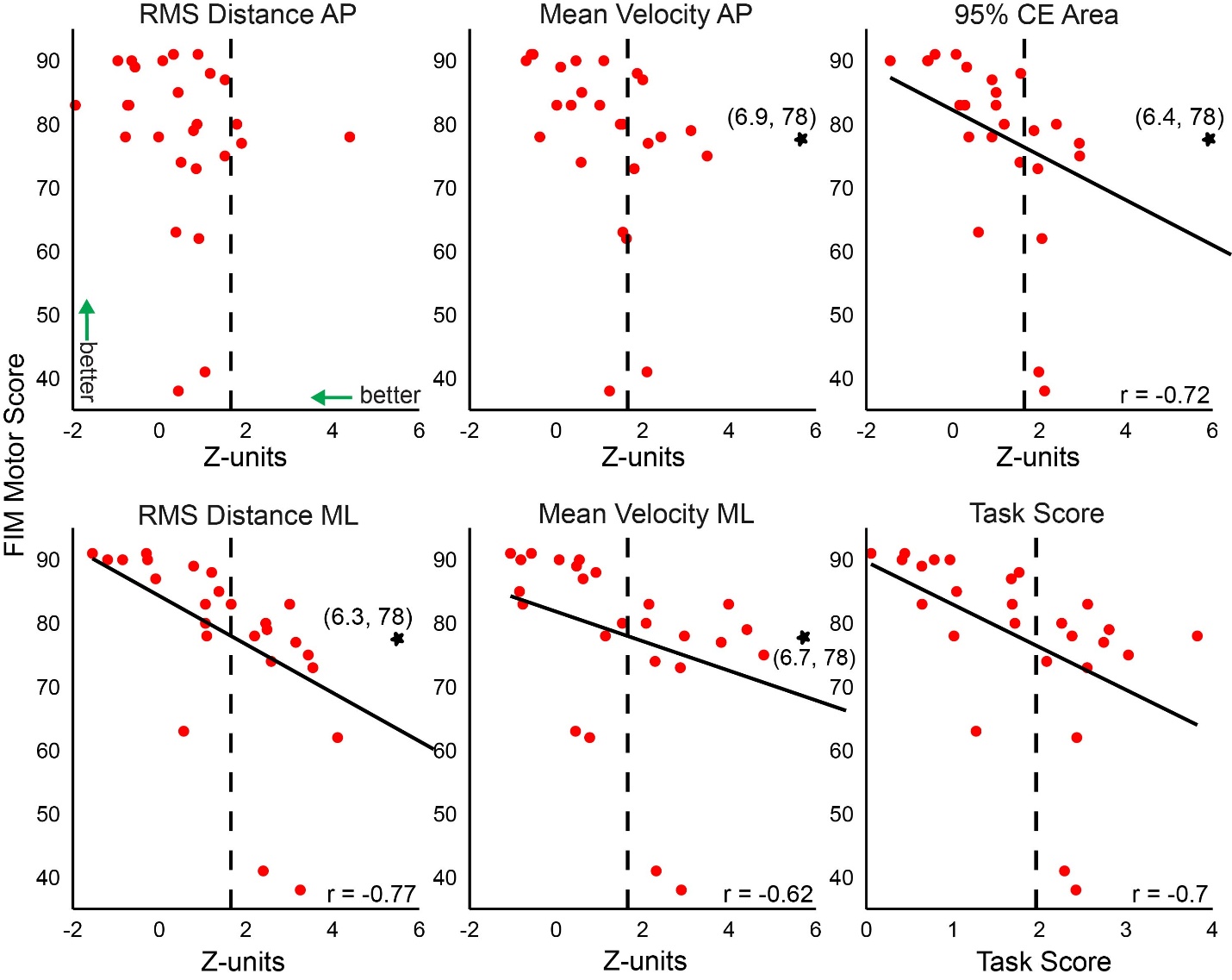
**Figure S6. Correlations between FIM-M scores and task parameters/Task Score.** Each red dot represents a participant with stroke. The dotted vertical line at Z = 1.64 for each discrete parameter represents the 95^th^ percentile of control performance, which was the threshold of impairment. A similar vertical dotted line at Task Score = 1.96 represents the same threshold for Task Score. One subject had Z-scores beyond the plotted range for several parameters and is indicated by a star on the relevant plots.

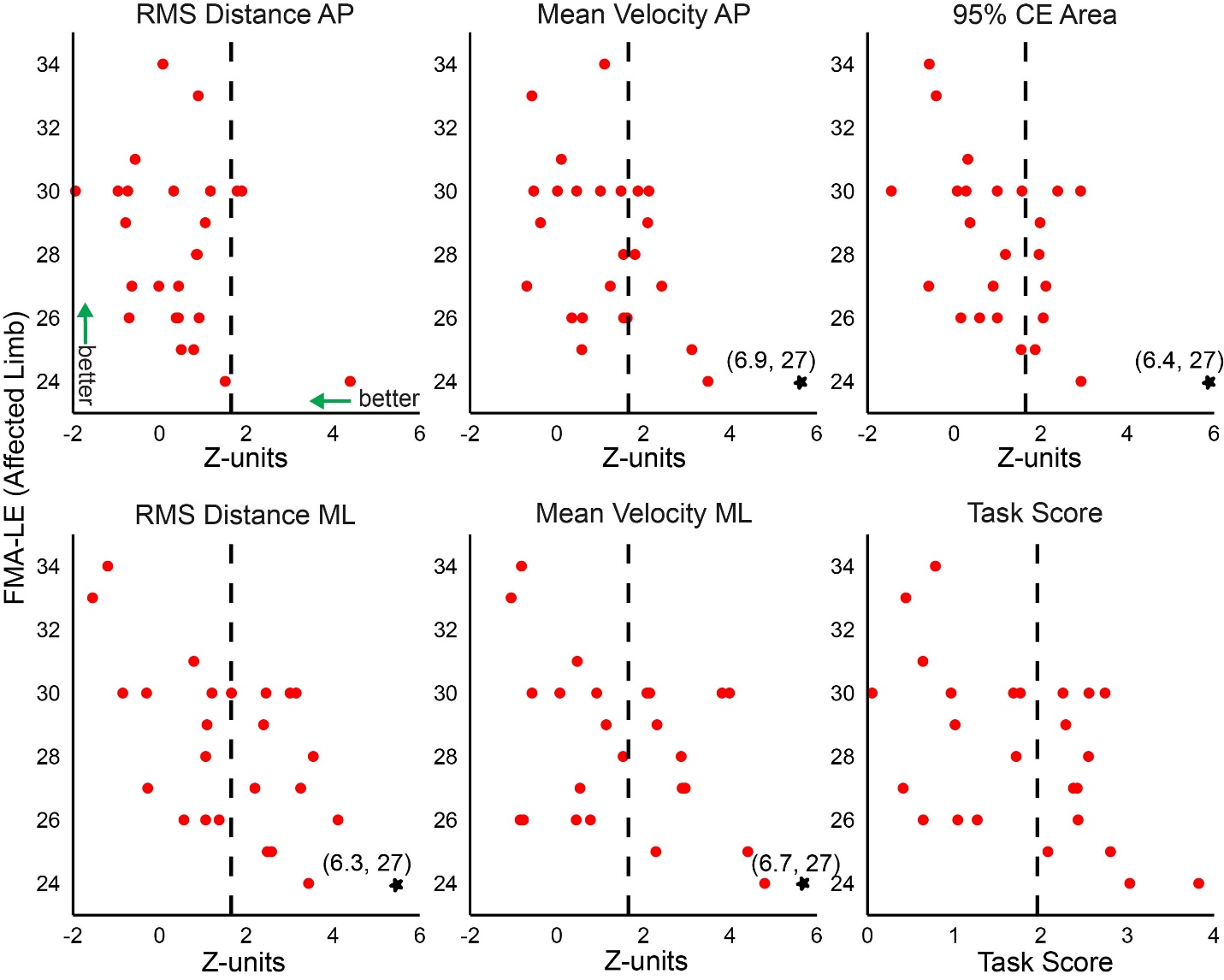
**Figure S7. Correlations between FMA-LE scores and task parameters/Task Score.** Each red dot represents a participant with stroke. The dotted vertical line at Z = 1.64 for each discrete parameter represents the 95^th^ percentile of control performance, which was the threshold of impairment. A similar vertical dotted line at Task Score = 1.96 represents the same threshold for Task Score. One subject had Z-scores beyond the plotted range for several parameters and is indicated by a star on the relevant plots.

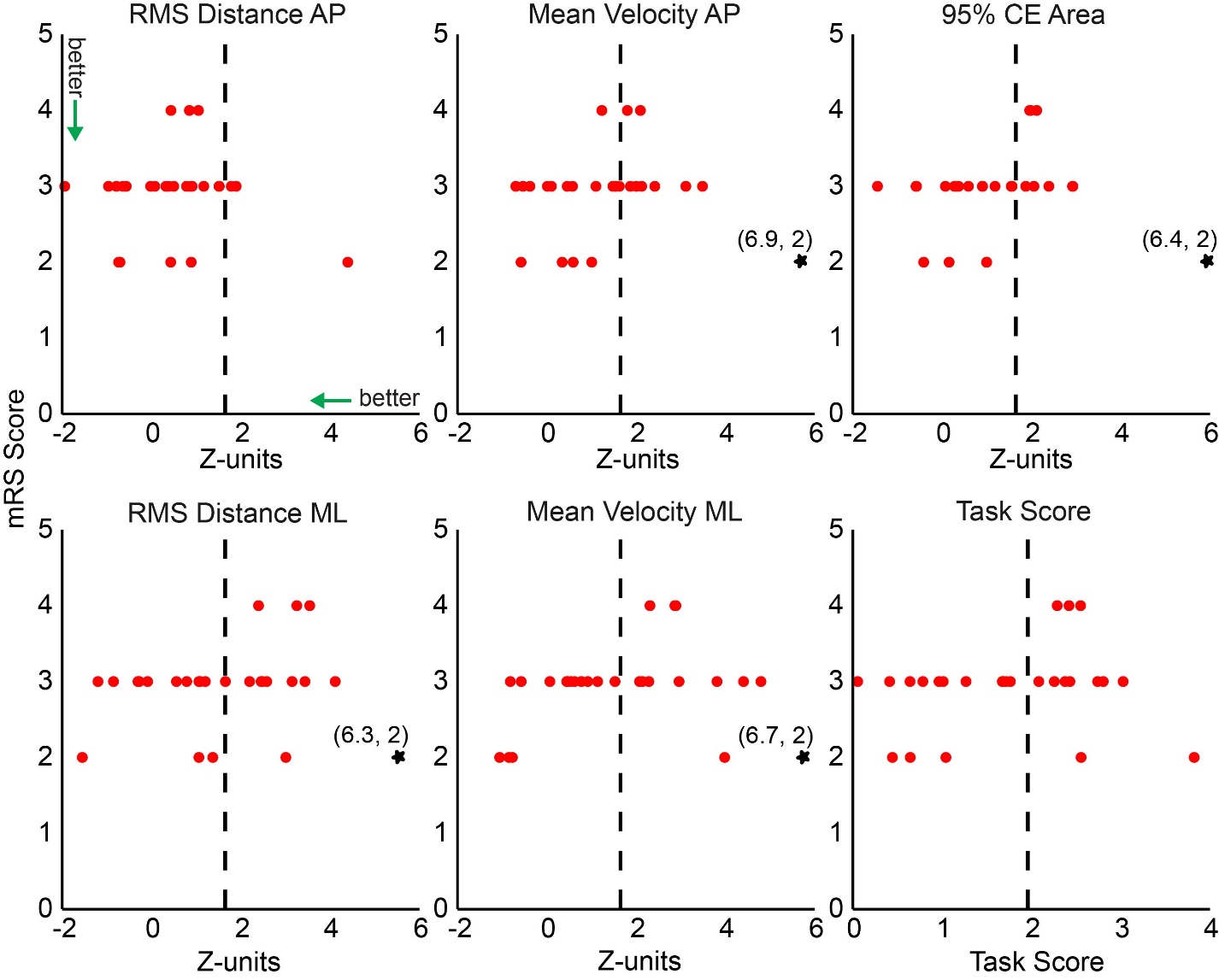
**Figure S8. Correlations between mRS scores and task parameters/Task Score.** Each red dot represents a participant with stroke. The dotted vertical line at Z = 1.64 for each discrete parameter represents the 95^th^ percentile of control performance, which was the threshold of impairment. A similar vertical dotted line at Task Score = 1.96 represents the same threshold for Task Score. One subject had Z-scores beyond the plotted range for several parameters and is indicated by a star on the relevant plots.

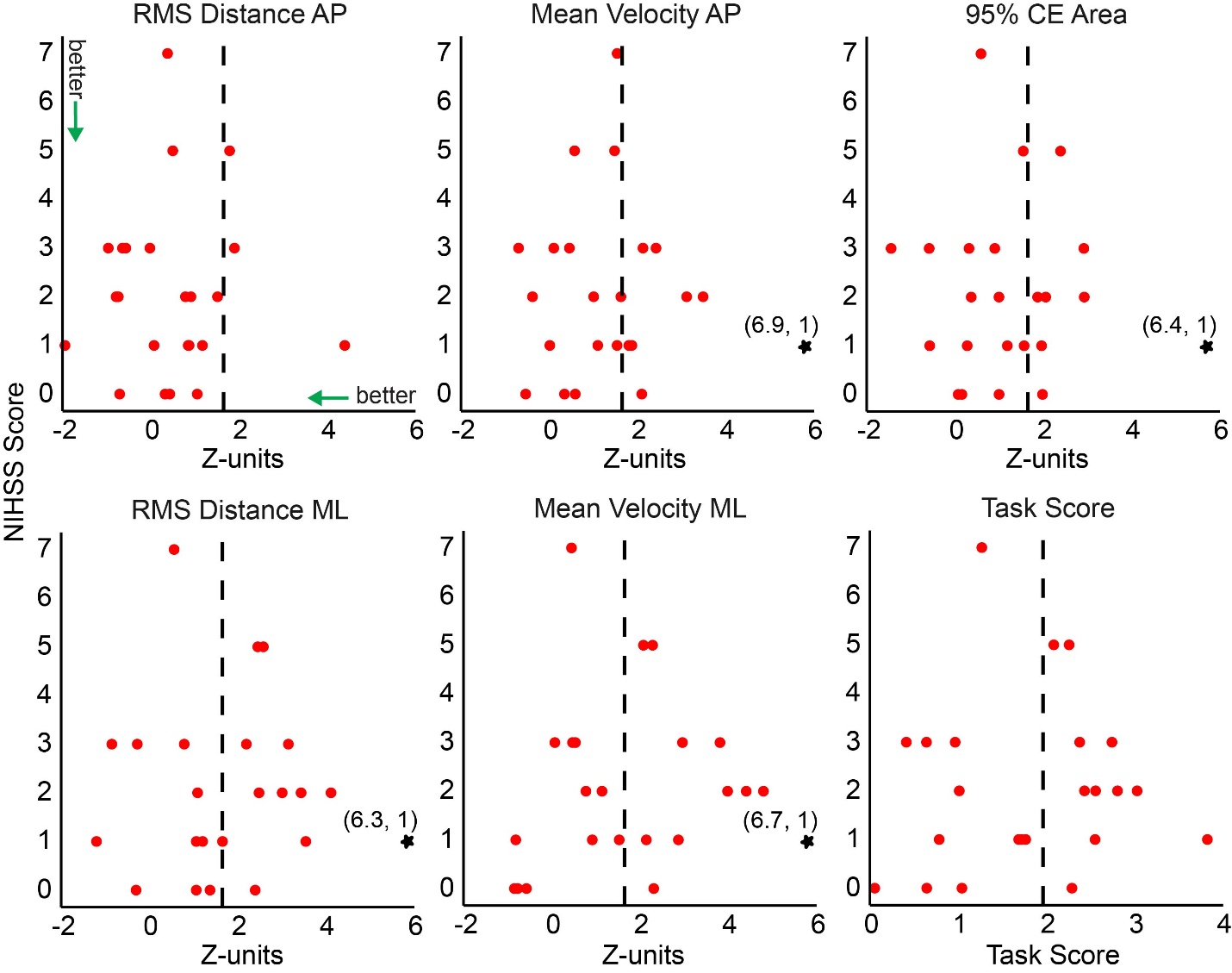
**Figure S9. Correlations between NIHSS scores and task parameters/Task Score.** Each red dot represents a participant with stroke. The dotted vertical line at Z = 1.64 for each discrete parameter represents the 95^th^ percentile of control performance, which was the threshold of impairment. A similar vertical dotted line at Task Score = 1.96 represents the same threshold for Task Score. One subject had Z-scores beyond the plotted range for several parameters and is indicated by a star on the relevant plots.
